# Differences in the cost of manufacturing pharmaceuticals in India for differently regulated markets: A comparative analysis

**DOI:** 10.1101/2025.04.16.25325941

**Authors:** Shireesh Ambhaikar, Shantaram Shenai, V Govindarajan, Pavan Gajare, P K Kulkarni, Shankar Suryanarayanan, Christopher Peterson, Lauren Schoukroun-Barnes, Meghana Inamdar, Sayan Paria, Giancarlo Francese, Murray Lumpkin, Jagdish Dore, Jean Christophe Rusatira, Charles Preston

## Abstract

**Background:** Ensuring access to affordable, high-quality pharmaceuticals is essential for global health. However, regulatory requirements vary significantly across markets, influencing manufacturing costs, drug prices, and access to quality medicines. This study examines cost variations in pharmaceutical manufacturing in India for products destined for three distinct regulatory markets: the United States (highly regulated), the World Health Organization (WHO) Prequalification (highly regulated), and India (regulatory stringency comparable to many Lower- and Middle-Income Countries). Understanding these cost differences provides valuable insights for regulators, policymakers, and procurers striving to balance affordability, quality, and accessibility.

**Methods:** The study utilizes a bottom-up costing approach to estimate manufacturing expenses for small-molecule generic drugs produced in India for different regulatory markets. Cost estimates are based on real-world industry data from Sidvim, a pharmaceutical manufacturing consultancy. The analysis distinguishes between capital expenditures (CAPEX) and operational expenditures (OPEX), comparing costs associated with active pharmaceutical ingredient (API) and finished dosage form (FDF) production. To isolate cost variations related to regulatory requirements, the study standardizes manufacturing conditions, including production scale, volumes, and facility utilization.

**Results:** The findings indicate substantial cost differences based on regulatory market. Manufacturing API and FDF for the Indian market incurs 43% lower CAPEX and 47% lower OPEX compared to the U.S. Key cost drivers include regulatory compliance, facility infrastructure, equipment standards, quality control, and personnel costs. Facilities serving highly regulated markets require stringent quality control, high-grade equipment, and frequent inspections, contributing to elevated costs. The study highlights the need for research on trade-offs between affordability and quality. Enhancing international collaboration and regulatory harmonization may facilitate access to high-quality, affordable medicines globally.

**Conclusion:** Cost disparities in pharmaceutical manufacturing underscore the challenge of balancing affordability with quality. Further research is needed to assess how these cost variations impact patient access, informing regulatory strategies and procurement policies across diverse market settings.

## Introduction

Ensuring access to affordable, safe, effective, and quality medicines is a cornerstone of global health and a critical component of achieving Universal Health Coverage (UHC).(1-4) There are many aspects to doing this, but one of the understudied facets is that drug manufacturers reduce the costs of manufacturing for different markets, which can improve affordability in the context of complex societal priorities and healthcare environments but little is known about these manufacturing differences, their drivers, and the effects on product safety, efficacy, and quality.(5, 6) Understanding this is crucial so that stakeholders—mainly National Regulatory Authorities (NRAs), as well as other relevant actors, such as national and global procurers, can assess implications and calibrate actions regarding drug regulation and procurement.

The cost of pharmaceutical manufacturing is influenced by several factors, including market regulatory requirements, production scale, production efficiency, the device’s cost (if applicable for combination products) and the cost of active pharmaceutical ingredients (APIs).(7, 8) Data from pharmaceutical companies have shown that manufacturing costs can constitute around 20-30% of total expenses inclusive of direct material costs for patent protected brand-name drugs and around 52% for generic drugs.(9) We also know these manufacturing costs can vary depending on the regulatory stringency of the market. For example, manufacturing for highly regulated markets like the United States involves more quality control measures which in turn increases manufacturing costs compared to less stringent markets such as those in low- and middle-income countries (LMICs).(9-11)

Although we know manufacturing costs can vary, there is a critical knowledge gap around the magnitude and drivers of the differences in costs of manufacturing for targeted markets.(9, 12). Various studies that focused on the extent and drivers of production have mainly focused on firm-specific factors such as the firm size, (13)the types of innovations, (6) income-level of the manufacturing country,(14) differences in prices of similar generics in different markets(15), and have all underscored the scarcity of detailed analyses on development and manufacturing cost variations for products destined for different markets.

Reducing manufacturing costs can be a strategy to address various factors in the system that push down pharmaceutical prices, such as the mix of health insurance coverage to out-of-pocket spending, government/purchaser expectations for affordable and lower-priced products and price controls.(16, 17) When these different manufacturing strategies are employed, along with their differential associated costs, it is important to keep in mind that companies will likely sell their less expensive-to-manufacture versions in lower regulatory environments, such as LMICs.

With this context in mind, our paper bridges the knowledge gap of drivers of different manufacturing costs for different markets by examining the costs of pharmaceuticals manufactured in India destined for the United States, the World Health Organization (WHO) prequalification program, and India. We chose India as the manufacturing base because it is a major supplier of generic drugs to the world, exporting to more than 200 countries. We chose the United States as a destination market because it is known for having one of the most well-regulated markets in the world, overseen by the Food and Drug Administration (FDA), and because it imports a significant percentage of its generic medicines from India.(9, 10) We chose WHO PQ as an intended market because it constitutes the United Nation’s mechanism for quality assurance of products purchased, and the program reviews a high percentage of products manufactured in India.(18, 19) We chose India as a destination market because it is governed by an emerging regulatory authority, and is likely tantamount to other LMIC destination markets.(20)

Thus, this study is intended to help regulators, as well as other stakeholders, such as national procurers, better understand drivers of the differences in the manufacturing of pharmaceuticals for intended markets. We discuss some limited implications of these findings but hope this work can spur additional studies in the area.

## Methods

### Design

The paper’s methods and results are based on industry and expert knowledge of the costs of manufacturing for different markets. The study data originated from internally available information within Sidvim LifeSciences Private Limited (Sidvim).(21) Sidvim is an India-based company that has been involved in various local and global pharmaceutical manufacturing initiatives(22), including helping design and establish more than 130 manufacturing and development facilities in India and other countries. Sidvim’s model is based on experience, which translates into more than 90% of its team of experts having at least 20 years of operating experience in the industry. To secure valid data, Sidvim used its own database of prices that factored in the local Indian context and consulted industry peers to ensure the accuracy and recency of internally available information. The estimation of manufacturing costs for different regulated markets was guided by an overarching set of assumptions (see Appendix C) and the framework shown in Figure 1. The costs generated by the framework are shown in the results. The paper does not analyze the implications for drug quality, the interplay with affordability, or go into detail about the needed regulatory strategies to address differences in manufacturing.

**Figure 1.**
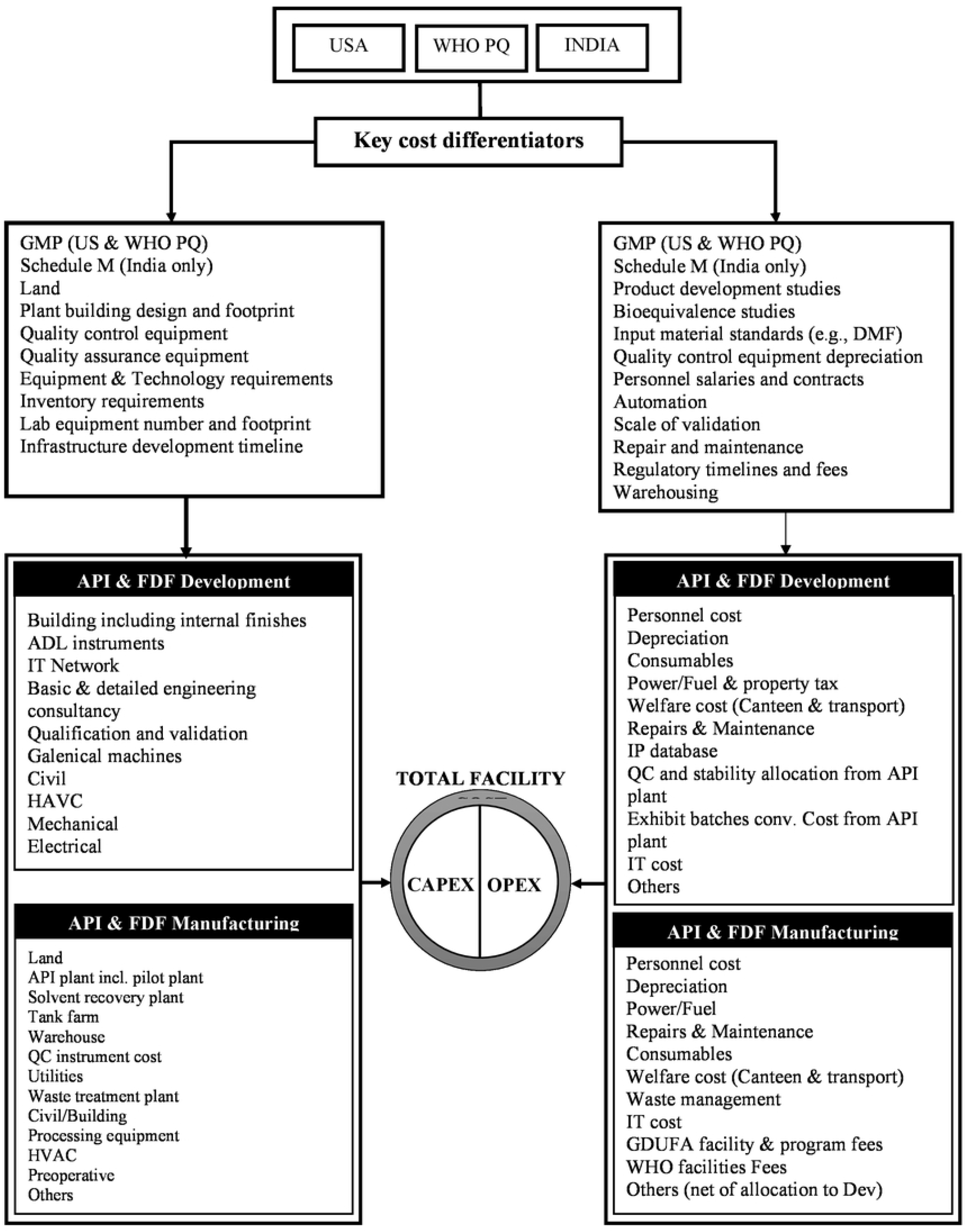
Conceptual framework for the estimation of total facility costs.

### Setting

To conduct this assessment, we estimated the costs of manufacturing small molecule generic drugs in three facilities based in India destined for three respective markets: the United States, WHO PQ, and the Indian market. The facilities, all assumed to be located in Aurangabad city, Maharashtra state, India, were analyzed to determine current fair-market compensation for services provided per manufacturing activity. The costs were estimated using a bottom-up approach for “greenfield plants” defined as a facility built with little-to-no existing infrastructure.(23) To ensure cost comparability across different facilities, factors such as production scale, volumes, and similar production time frames were held constant. Case study facilities considered were differentiated based on product destination market-specific regulatory requirements, which have varying inputs such as product development and clinical/bioequivalence studies, equipment data requirements (e.g., design and performance qualification), regulatory approval timelines, and commercial needs such as warehousing capacity. The capacity and utilization levels were standardized across case study facilities to eliminate the impact of economies of scale. However, the costs of the direct material inputs (i.e. intermediates, API and excipients, packaging material etc.) were not included as those are molecule-specific and not related to manufacturing conversion costs which is the focus of this analysis.

Further, the costs for US-destined products were estimated based on FDA requirements, including those that adhere to the International Council for Harmonisation (ICH) of Technical Requirements for Pharmaceuticals for Human Use Good Manufacturing Practices (GMPs).(24, 25) Similarly, WHO PQ also generally follows ICH GMP (24-26) In the case of India, despite some variations across states, the requirements and enforcement frameworks are similar to those in many LMICs, which, like India, have national regulatory agencies at level 1 or 2 for oversight of medicines, as measured on WHO’s Global Benchmarking Tool.(19) These maturity levels indicate that the regulatory systems are still evolving, with limited or basic regulatory functions in place, and significant improvements are needed to ensure effective regulation.(27)

### Conceptual framework

We used a well-established conceptual framework to estimate manufacturing costs, composed of two components, namely Capital Expenditures (CAPEX) and Operational Expenditures (OPEX).(28-30) This financial framework has been previously used in pharmaceutical manufacturing studies to categorize and manage organizational expenses. (28-30)The sum of CAPEX and OPEX costs yielded the total costs of the facility which were compared to estimate the differences in manufacturing costs for each of the three intended markets. We termed the total costs for each facility to make products for the three destination markets “total facility cost.” The three facilities were assumed to produce several generic oral solid tablets of moderate complexity commonly used in clinical care situations. This assumption was made because oral solid pharmaceuticals are simpler to produce and provide greater chemical and physical stability, making costing estimations more accurate.(31) These tablets were omeprazole, amlodipine, atorvastatin, clopidogrel, sertraline, and linezolid. However, the costs of the direct material inputs (i.e. intermediates, API and excipients, device if applicable, packaging material etc.) were not included as those are molecule-specific and not related to manufacturing conversion costs.

For each destination market, the facility costs were estimated based on regulatory guidelines, with areas of focus, such as Good Manufacturing Practices (GMP) at the manufacturing site and regulatory documentation for approval. The costs were estimated for a greenfield site making both active pharmaceutical ingredient (API) and final dosage forms (FDF). The costs for API and FDF were split into two categories: 1) CAPEX costs for setting up the plant and pre-operative expenses and 2) OPEX. Each of the two categories had one block for development and another for manufacturing. The total facility costs result from CAPEX and OPEX as shown in Figure 1.

The CAPEX costs are defined as investment costs attributable to the facility development and are comprised of two major categories: API and FDF development and API and FDF manufacturing. CAPEX costs are further subdivided into investments in items like IT, basic and detailed engineering, land, API plant, solvent recovery plant, etc. per Figure 1. CAPEX component costs result from a summation of associated investment costs, fees, and equipment requirements to establish and continually run the facility. The details of CAPEX components and methods of calculation are shown in Appendix A.

OPEX costs are defined as costs associated with operating and maintaining the proposed facility. There are two major categories, API and FDF development and API and FDF manufacturing. OPEX is further subdivided into personnel, depreciation, power, repairs and maintenance, consumables, etc. across these two categories, per Figure 1. Each OPEX component is an associated cost compiled from operating various aspects of the facility, maintaining equipment and fees associated with released drugs in regulated markets. To account for each operational cost, the proposed facility is subdivided into cost centers. The cost centers encompass a functional department with the facility, and the details for each center are provided below. The details of OPEX components and methods of calculation are shown in Appendix B.

We used several assumptions to make this exercise simpler for the analysis, which are listed in Appendix C. All calculations were performed with Microsoft Excel 2015.(32)

## Results

The differences in the costs of Indian-made products for different regulatory markets are described below (x1000$). The costs are presented by category and block. However, as the US and WHO PQ costs are so similar, we have eliminated the WHO comparison in the descriptions below, but it is included in the tables.

### 1. Active Pharmaceutical Ingredient

The total API CAPEX and OPEX are 32-37% lower for India-destined compared to US-destined products. (Table 1) The differences in API CAPEX and OPEX between the US and WHO PQ-destined products are at around 1-2%, respectively.

**Table 1.** Average costs for Active Pharmaceutical Ingredient development and manufacturing.

| | US-destined<br>products<br>(Ref.) | WHO<br>PQ-destined<br>products | Difference<br>US - WHO<br>PQ-destined<br>Diff % | India-destined<br>products<br>(x1000\$) | Difference<br>US - India<br>destined<br>Diff % |
| --- | --- | --- | --- | --- | --- |
| **Asset Block** | **(x1000\$)** | **(x1000\$)** | | | |
| **CAPEX** |  |  |  |  |  |
| **API Development** |  |  |  |  |  |
| Building including internal finishes | 571 | 571 | 0.0% | 286 | 49.9% |
| ADL instruments | 1,624 | 1,414 | 12.9% | 349 | 78.5% |
| IT Network | 45 | 40 | 11.1% | 28 | 37.8% |
| Basic and detailed eng. Consultancy | 20 | 20 | 0.0% | 10 | 50.0% |
| Qualification and validation | 20 | 18 | 10.0% | 4 | 80.0% |
| **Sub-total** | **2,280** | **2,064** | **9.5%** | **677** | **70.3%** |
| **API manufacturing** |  |  |  |  |  |
| Land | 2,302 | 2,302 | 0.0% | 2,021 | 12.2% |
| API plant including pilot plant | 8,631 | 8,631 | 0.0% | 4,997 | 42.1% |
| Solvent recovery plant | 1,298 | 1,298 | 0.0% | 1,298 | 0.0% |
| Tank farm | 481 | 481 | 0.0% | 279 | 42.0% |
| Warehouse | 986 | 986 | 0.0% | 828 | 16.0% |
| Quality control | 1,490 | 1,490 | 0.0% | 727 | 51.2% |
| Utilities | 2,038 | 2,038 | 0.0% | 1,967 | 3.5% |
| Waste treatment plant | 1,107 | 1,107 | 0.0% | 1,107 | 0.0% |
| Preoperative | 1,806 | 1,806 | 0.0% | 995 | 44.9% |
| Others | 1,866 | 1,866 | 0.0% | 1,520 | 18.5% |
| **Sub-total** | **22,004** | **22,004** | **0.0%** | **15,739** | **28.5%** |
| **Total CAPEX** | **24,284** | **24,068** | **0.9%** | **16,416** | **32.4%** |
| **OPEX** |  |  |  |  |  |
| **API development** |  |  |  |  |  |
| Personnel cost | 665 | 594 | 10.7% | 241 | 63.8% |
| Depreciation | 402 | 353 | 12.2% | 66 | 83.6% |
| Consumables | 181 | 181 | 0.0% | 32 | 82.3% |
| Welfare cost (Canteen & transport) | 37 | 31 | 16.2% | 21 | 43.2% |
| Repairs & Maintenance | 20 | 19 | 5.0% | 10 | 50.0% |
| IP database | 24 | 24 | 0.0% | 2 | 91.7% |
| QC and stability allocation from API plant | 116 | 116 | 0.0% | 23 | 80.2% |
| Exhibit batches conv. Cost from API plant | 292 | 292 | 0.0% | 59 | 79.8% |
| IT cost | 14 | 13 | 7.1% | 9 | 35.7% |
| Others | 178 | 173 | 2.8% | 96 | 46.1% |
| **Sub-total** | **1,930** | **1,796** | **6.9%** | **559** | **71.0%** |
| **API manufacturing** |  |  |  |  |  |
| Personnel cost | 2,102 | 2,102 | 0.0% | 1,112 | 47.1% |
| Depreciation | 1,508 | 1,508 | 0.0% | 979 | 35.1% |
| Power/Fuel | 1,533 | 1,533 | 0.0% | 1,713 | -11.7% |
| Repairs & Maintenance | 522 | 522 | 0.0% | 382 | 26.8% |
| Consumables | 399 | 399 | 0.0% | 270 | 32.3% |
| Welfare cost (Canteen & transport) | 239 | 239 | 0.0% | 158 | 33.9% |
| Waste management | 96 | 96 | 0.0% | 96 | 0.0% |
| IT cost | 58 | 58 | 0.0% | 56 | 3.4% |
| GDUFA facility & program fees | 53 | 0 | 100.0% | 0 | 100.0% |
| WHO facilities Fees |  | 54 | n/a |  | n/a |
| Others (net of allocation to Dev) | 179 | 171 | 4.5% | 123 | 31.3% |
| **Sub-total** | **6,690** | **6,682** | **0.1%** | **4,888** | **26.9%** |
| **Total OPEX** | **8,620** | **8,478** | **1.6%** | **5,447** | **36.8%** |

#### a. Capital Expenditure for API development

Overall, US-destined products have the highest total CAPEX at $2,280K. In contrast, India-destined costs are $677K, corresponding to a reduction of 70.3% relative to the US-destined products. The building costs for India-destined products are 49.9% lower than the US. The most notable differences are observed in the costs of Analytical Development Laboratory (ADL) equipment, qualification and validation, with India-destined costing 78.5% and 80.0% less, respectively, compared to the US.

#### b. Capital Expenditure for API Manufacturing

The CAPEX for API manufacturing for the Indian market is around 28.5% lower than manufacturing for the other markets. For land acquisition, facilities manufacturing for India require around $2,021K (12.2% lower than the US-destined manufacturing). The expenditures on the API plants including the pilot plants to manufacture for India is $4,997K (42.1% lower than compared to the US). Other notable differences include quality control costs for the US market at $1,490K, while the costs for India-destined manufacturing are at $727K, a 51.2% reduction.

#### c. Operational Expenses for API Development

Personnel costs for the US-destined products are $665K and $241K for India-destined products, a 63.8% reduction compared to the US-destined products. Other categories, such as consumables and welfare costs, show varied reductions across markets. For instance, welfare costs, comprising food and transport, for facilities developing APIs for the Indian market are 43.2% lower than in those developing APIs for the US market.

#### d. Operational Expenses for API Manufacturing

Personnel costs for the US-destined products are $2,102K, while India-destined products are at $1,112K, which is 47.1% lower compared to US-destined products. Other notable differences exist in costs for repairs and maintenance, consumables, and welfare. For example, repairs and maintenance cost $382K for India-destined APIs, which is 26.8% lower than for the US-destined APIs. Consumables and welfare costs are reduced by 32.3% and 33.9%, respectively, when comparing India-destined to US-destined APIs.

### 2. Finished Dosage Form Costs Assessment

The overall FDF costs comprise the capital expenditures for FDF development and FDF manufacturing together with the operational expenditures for FDF development and FDF manufacturing. (Table 2) The FDF development CAPEX and OPEX are respectively 68.6% and 65.5% lower for facilities developing products destined for India compared to those facilities producing products for the US. The FDF manufacturing CAPEX and OPEX are respectively 52.4% and 53.1% lower for products destined for India compared to those destined for the US.

**Table 2.**
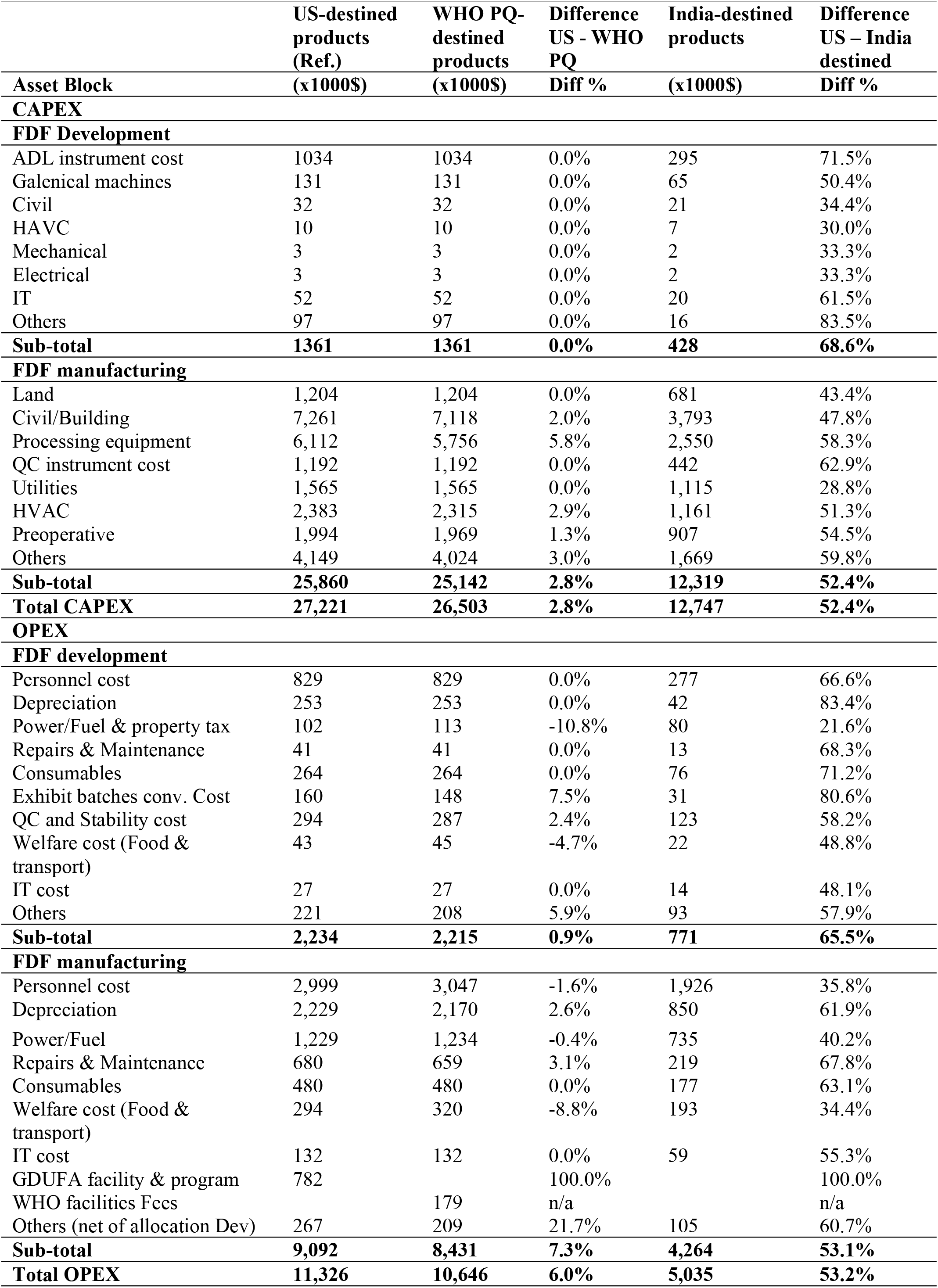
Average costs for Final Dosage Form Development and Manufacturing.

#### a. Capital Expenditure for FDF development

The most substantial cost difference is observed in the ADL equipment cost, where the products destined for the US are at $1,034K. In contrast, India-destined products show a lower cost of $295K, reflecting a 71.5% reduction. While for the US-destined products, the costs are around $131K for galenical machines essential to produce solid and semi-solid dosage forms, for India-destined products the costs are just $65K, which is 50.4% lower than the other regions. The information technology cost for India-destined products is $20K, showing a reduction of 61.5%.

#### b. Capital Expenditure for FDF Manufacturing

For land acquisition, US-destined manufacturing costs are around $1,204K compared to $681K for India-destined products, which is 43.4% lower. Civil and building costs exhibit similar trends with India-destined products showing a cost of $3,793K, 47.8% lower than costs for the US-destined products. For the US-destined products, processing equipment costs $6,112K. For India-destined products, however, only $2,550K for processing equipment is incurred, a 58.3% reduction. Similarly, Quality Control (QC) instrument costs for India-destined products are $442K, 62.9% lower than the $1,192K reported for the US.

#### c. Operational Expenses for FDF Development

Personnel costs are consistent with WHO PQ for the US at around $829K. However, India-destined products show a personnel cost of $277K, which is 66.6% less than for the US-destined products. Similarly, consumable costs for India-destined products stand at $76K, which is 71.2% lower than the $264K reported for products destined for the other markets.

#### d. Operational Expenses for FDF Manufacturing

For products destined for the US market, personnel cost is $2,999K. In contrast, for products destined for the Indian market, the personnel cost is $1,926K, which is 35.8% lower than for products destined for the US market. Repairs and maintenance costs are 67.8% lower for India-destined products compared to the US-destined products. Consumable costs are identical for US and WHO PQ products, however, for the India-destined products a saving of 63.1% was recorded.

### 3. Total API and FDF CAPEX and OPEX Comparison

The overall costs for API and FDF development and manufacturing are displayed in Figure 2. The API and FDF total CAPEXs are the additions of respective development and manufacturing CAPEXs. Similarly, API and FDF OPEXs are the additions of respective development and manufacturing OPEXs. The API and FDF costs for the US market show a total API FDF CAPEX of $51,505K for US and $29,163K India-destined products, which is 43.4% lower. The total API FDF OPEX is $10,482K for India-destined products, which is 47.4% lower than $19,946 for US-destined products.

**Figure 2.**
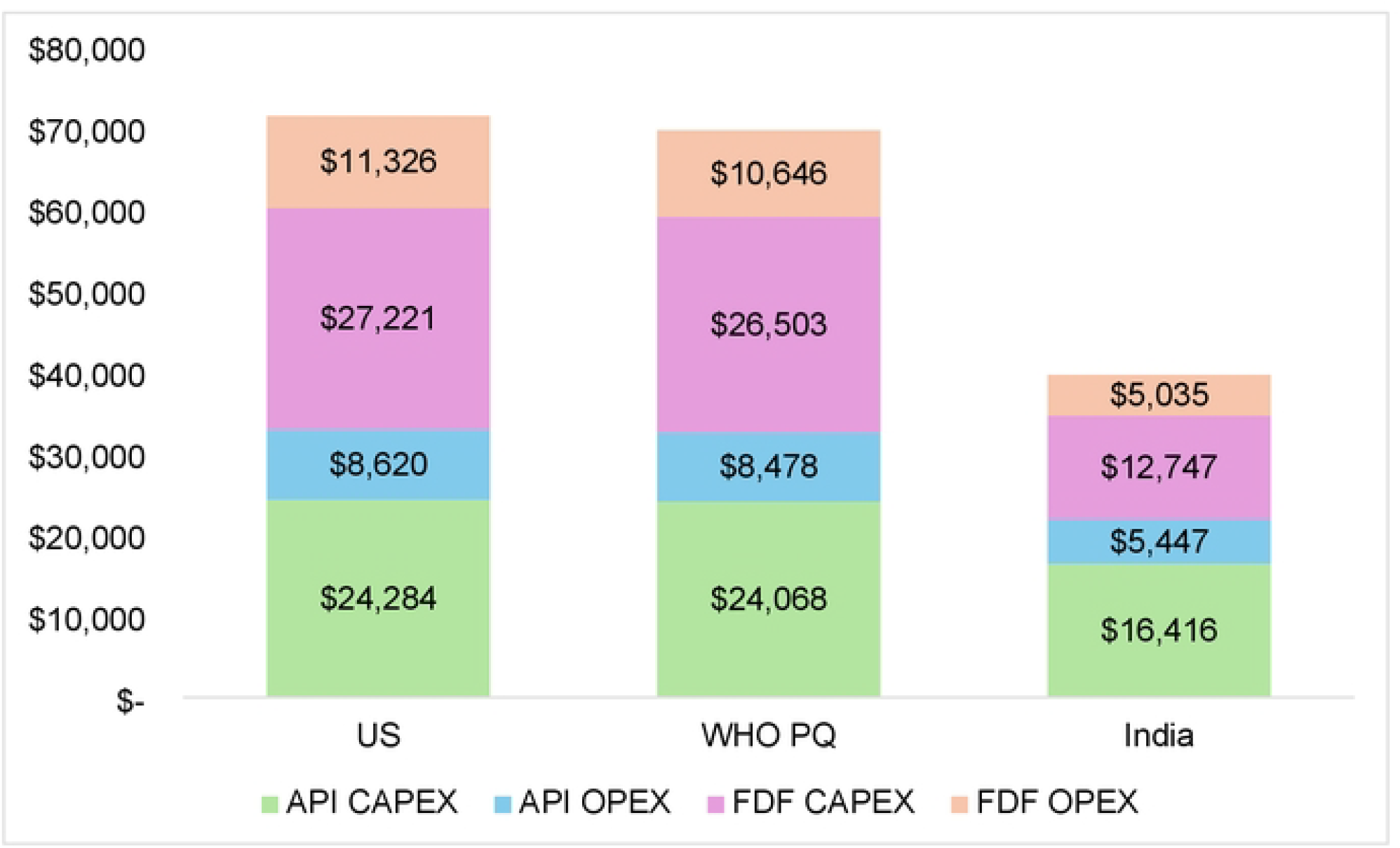
Comparison of API and FDF costs of pharmaceutical products in India based on the destined regulatory market. Note: CAPEX and OPEX include development and manufacturing totals.

## Discussion

This study provides unique comparative insights into the differences in manufacturing costs for Indian-made medicines depending on the destination market – the US, WHO PQ, or India itself. This exercise suggests that production costs for less regulated markets, such as India (and by extension other LMICs, for example, that have not reached maturity level 3 for medicines(27)), can be significantly cheaper, up to 43% lower in capital expenses and 47% in operational expenses, compared to products made in India for highly regulated markets.

Multiple factors appear to be driving the expenditure differences. These stem from land size and equipment costs. For example, the facilities manufacturing for the Indian market have smaller land sizes, utilize less expensive equipment, such as lower-grade stainless steel or glass linings, and have less complex closed systems. These facilities also employ less preventive maintenance, calibration, and automation, with minimal reliance on real-time data acquisition systems. In contrast, similar to what has been documented in the literature, facilities manufacturing for mature markets incur higher costs due to the use of more advanced equipment and greater automation. (9-11)

The differences in operational costs have multiple drivers as well. Facilities manufacturing for the Indian market tend to use less complex equipment and operate with less preventive maintenance, calibration, and automation, and tend to rely on just-in-time (JIT) manufacturing and minimal warehousing. Additionally, smaller facilities with fewer cleaning requirements and shorter downtimes further reduce costs. India-destined products also leverage a lower-paid workforce, compared to workers at facilities manufacturing for higher regulated markets, with a bigger proportion of contract labor. In contrast, manufacturing for highly regulated mature markets uses more intensive operational processes, including stricter cleaning protocols, greater reliance on automation, and higher real-time data acquisition standards, leading to significantly higher operational costs. Moreover, facilities manufacturing for the US incur additional expenses due to stringent documentation, traceability, frequent inspections, quality control costs, and higher regulatory fees.

These findings suggest there is an economic rationale to manufacturing pharmaceuticals, wherein manufacturers may prioritize different cost models depending on the regulatory standards and enforcement variabilities of the target market, and in the context of other factors like pricing pressures. These findings corroborate with existing literature and put a spotlight on the relationships between aims such as affordability and quality. (7, 8) (15) While products destined for LMICs may be less expensive to make, and theoretically more affordable, it also raises the issue of potential discrepancies in quality, where there are differences in workforce, equipment maintenance, quality control costs, and inspections, to name a few. More research is needed to identify any trade-offs between affordability and quality.

The study also has implications for regulatory reliance on reference NRAs, which is an important strategy for LMIC regulators to accomplish regulatory work with fewer resources.(33) Regulators and procurers must understand that more expensively manufactured products are typically made for higher regulated markets. When companies want to sell, for example, their “FDA approved” versions in an LMIC market, with less regulation, regulators and procures must check through version verification and market monitoring that they are getting, and will continue to receive, the exact same product version approved by the reference regulator (e.g. same manufacturing plant, same processing and controls, etc.).

This study has important limitations. While it uses real-world data to compare production costs for different regulatory environments, the focus on generic oral solid tablets limits the generalizability of the findings to other pharmaceutical forms. Additionally, the analysis assumes ideal conditions, such as a greenfield plant setup in a single location in India, which may not capture the complexities and variabilities of real-world production environments across different countries. The study focuses on differences in costs of production but does not directly assess affordability or quality differences between products produced for the different markets. Further, the estimates can be affected by rapid changes in foreign exchange rates and other changes in the pharmaceutical manufacturing industry. Lastly, the study relies on an expert group to model the costs of manufacturing. It is possible that other groups might use different prices and assumptions.

## Conclusion

This study showed substantial variations in pharmaceutical manufacturing costs when products made in India are destined for markets with different levels of regulatory oversight. Both capital expenses and operational expenses were substantially higher for the products manufactured for highly regulated markets like the US and WHO PQ than for India, the latter of which is similar to other LMIC destinations. These differences in the costs of manufacturing drugs for different markets, however, raise important questions, including any tradeoffs between affordability and product quality. Because there are substantive and material differences in manufacturing, it is important for regulators and procurers to understand these differences, as well as develop strategies to mitigate any unwanted effects. Lastly, while this study highlights critical patterns in manufacturing costs and the effect of regulatory requirements and enforcement variabilities, the broader implications, particularly in terms of patient access to affordable, safe, effective, quality pharmaceuticals warrant further investigation. The study lays a foundation for others to investigate these crucial aspects further. Such inquiries are necessary to inform national regulatory strategies and pharmaceutical procurement policies across diverse regulatory contexts.

## Data Availability

The data used for this study can be obtained from the corresponding author upon reasonable request.

## Supporting information

S1 Text. Capital Expenditures (CAPEX) Components used for Calculation

S2 Text. Operational Expenditures (OPEX) Components used for Calculation

S3 Text. List of Assumptions

